# A Diagnostic Blind Spot: Deep intronic SVA_E Insertion identified as the most Common Pathogenic Variant Associated with Canavan Disease

**DOI:** 10.1101/2024.12.18.24318806

**Authors:** Carlos A. Dominguez Gonzalez, Katrina M. Bell, Ramakrishnan Rajagopalan, Michelle G. de Silva, Aída Lemes, Cristina Zabala, Florencia Pérez, Alfredo Cerisola, Arastoo Vossough, Matthew T. Whitehead, Chloe Cunningham, Natasha J. Brown, Rebecca Quin, Cas Simons, Thomas Conway, Eloise Uebergang, Rocio Rius, Meutia A. Kumaheri, Emma Kotes, Ananya Vohra, Miranda PG Zalusky, Zachary B. Anderson, Sophie HR Storz, Sydney A. Ward, Joy Goffena, Jonas A. Gustafson, Susan M. White, Adeline Vanderver, Danny E. Miller

## Abstract

Canavan disease (CD) is a neurodegenerative disorder caused by biallelic disease-causing variants in the *ASPA* gene. Here, we utilized long-read sequencing (LRS) to investigate eight individuals clinically diagnosed with Canavan disease but without definitive genetic diagnoses. Our analyses identified a recurring previously unreported intronic SVA_E retrotransposon insertion within *ASPA* in all eight individuals. Surprisingly, the frequency of this variant in population databases suggests it is the most common pathogenic variant in *ASPA* and should be evaluated in diagnostic testing and carrier screening for CD. Additionally, this finding has implications for the broader rare disease community, as it highlights a substantial blind spot in standard short-read diagnostic pipelines, which historically have missed or overlooked these types of insertions. This discovery highlights the power of emerging technologies, such as LRS and RNA-sequencing (RNA-seq), to bring new classes of variants into diagnostic utility for genetic disorders like CD.

## Main Text

Canavan disease (CD) (OMIM# 271900) is an autosomal recessive disorder caused by biallelic loss-of-function variants in the *ASPA* gene. Children affected with CD present with progressive and irreversible decline of previously acquired motor and cognitive milestones. Symptoms typically appear following normal development during the first months of life, including macrocephaly, hypotonia, loss of muscle control, feeding difficulties, developmental delay (including motor and verbal skills), optic atrophy, and seizures. Clinical severity and disease progression are likely associated with the enzyme’s residual activity and conformational stability^1^. In addition to suggestive clinical findings, the diagnosis is established by elevated N-acetylaspartic acid (NAA) in urine through gas chromatography-mass spectrometry or in the brain through proton magnetic resonance spectroscopy^2^.

The *ASPA* gene is located on chromosome 17p13.2, contains six exons, and is ∼30 kb long. Although the Ashkenazi Jewish population has a higher incidence of CD mainly due to the founder *ASPA* variants p.Glu285Ala and p.Tyr231Ter^3^, individuals from all ancestry groups can be affected, and over 100 disease-causing variants have been submitted to variant databases such as Leiden Open Variation Database (LOVD), ClinVar, and HGMD. These variants include missense/nonsense, splicing, deletions, and insertions. In the context of emerging therapeutic approaches in CD (NCT04833907, NCT04998396), it is becoming increasingly important to define molecular causes in addition to biochemical testing, as genetic confirmation may be a prerequisite to participating in clinical trials or future approved therapies.

Here, we report eight individuals from seven families **(Fig. 1a)** with confirmed CD diagnoses based on clinical, biochemical, and neuroimaging evidence **(Fig. 1b, c)** in whom genetic testing, including the use of short-read genome sequencing (srGS), did not identify disease-causing variants on one or both alleles of the *ASPA* gene. The reported SVA_E insertion was identified independently by two groups simultaneously. One team performed targeted long-read sequencing (LRS) of *ASPA* and a panel of genes associated with neurodegenerative disorders in infancy on individual FI:1. Filtering for single nucleotide polymorphisms and SVs with allele frequencies less than 1% in *ASPA* revealed a single homozygous ∼2,600 bp SVA_E insertion in intron 4 of 5 (NM_000049.4) **(Fig. 2a)**. Separately, a second team performed manual analysis of srGS data in IGV of data from individual FVI:1, who had a single disease-causing variant identified by diagnostic testing. The candidate insertion was detected by identifying and analyzing the discordant read pairs and the soft-clipped sequence present on either side of the target site duplication, which matched the SVA_E sequence **(Fig. S1)**. Subsequent LRS of individual FVI:1 confirmed an SVA_E insertion. Once identified, the insertion was found in six additional individuals who had biochemical and clinical evidence of disease but did not have a confirmed genetic diagnosis of CD. The insertion was found to be homozygous in all individuals with no candidate variant previously identified by prior clinical testing (FI:1, FII:1, FIII:1) and in trans with a known pathogenic variant in all individuals with a single candidate variant identified by standard clinical testing (FIV:1, FIV:2, FV:1, FVI:1, FVII:1). Insertions were confirmed by LRS in the probands and their family members **(Fig. S2)**.

**Fig. 1.**
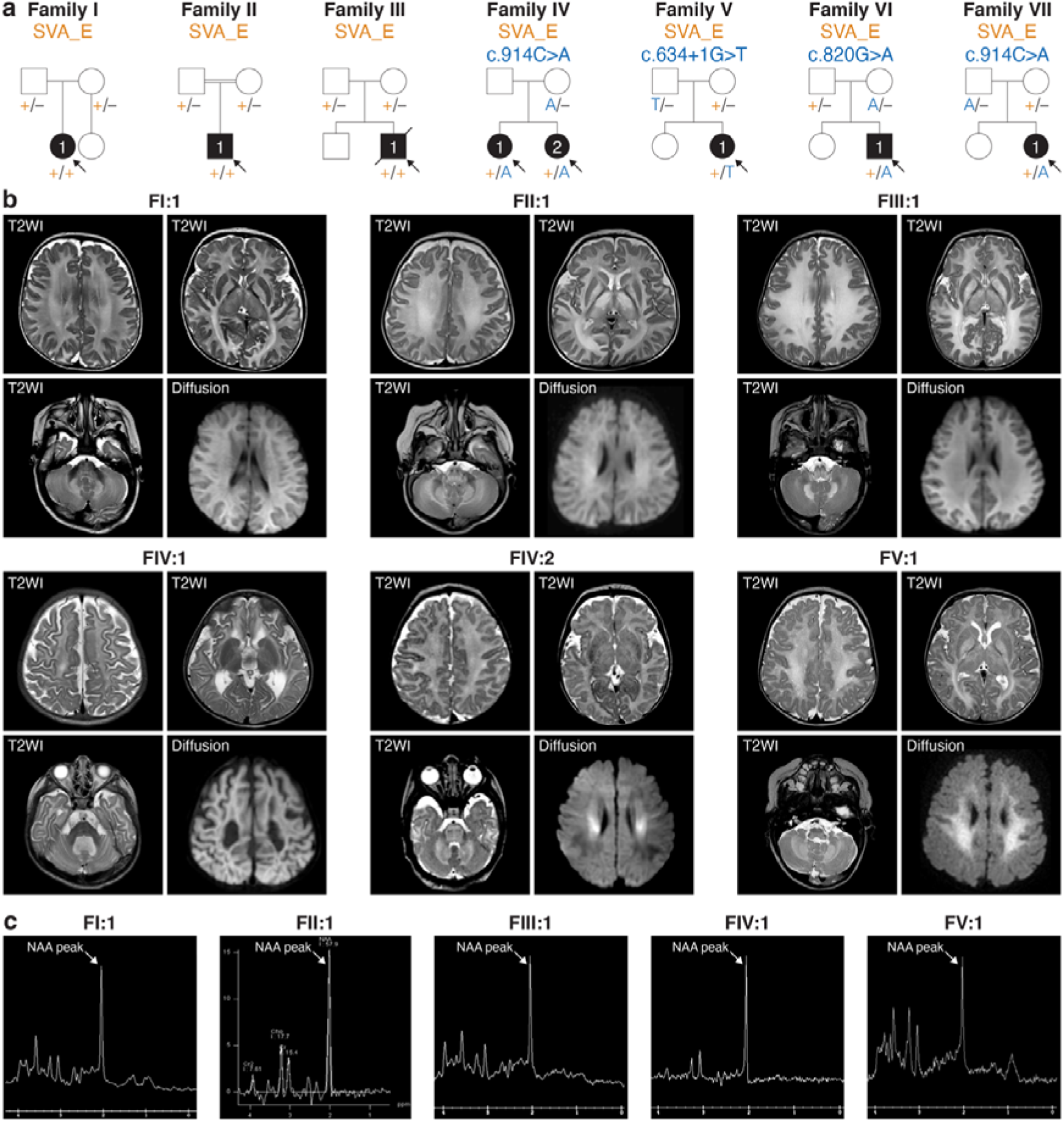
Pedigrees and MRI images of individuals with Canavan disease. **a,** Pedigrees showing segregation of the SVA_E insertion (+) in the *ASPA* gene. In individuals FI:1, FII:1, and FIII:1, the SVA_E insertion is homozygous. Among the other affected individuals, the insertion is in trans with the previously identified pathogenic variant. **b,** Brain MRI images corresponding to each patient (except FVII:1 in whom MRI was not performed, and FVI:1 whose neuroimaging was not available) grouped in panels of four; axial T2-weighted images through the centrum semiovale (top left), basal ganglia (top right), and posterior fossa (bottom left) and axial DWI through the centrum semiovale (bottom right). There is symmetric diffuse white matter signal abnormality with variable areas of reduced diffusivity, notably involving the globus pallidus, thalamus, and dentate nuclei. There is relative sparing of the corpus striatum, corpus callosum, and small focal regions of the posterior limb of the internal capsule. These imaging features are consistent with Canavan disease. **c,** Brain MR spectroscopy in each patient (except FIV:2 and FVII:1, in whom MRS was not performed, and FVI:1, whose neuroimaging was unavailable) reveal elevated NAA at 3 ppm, characteristic of Canavan disease.

**Fig. 2.**
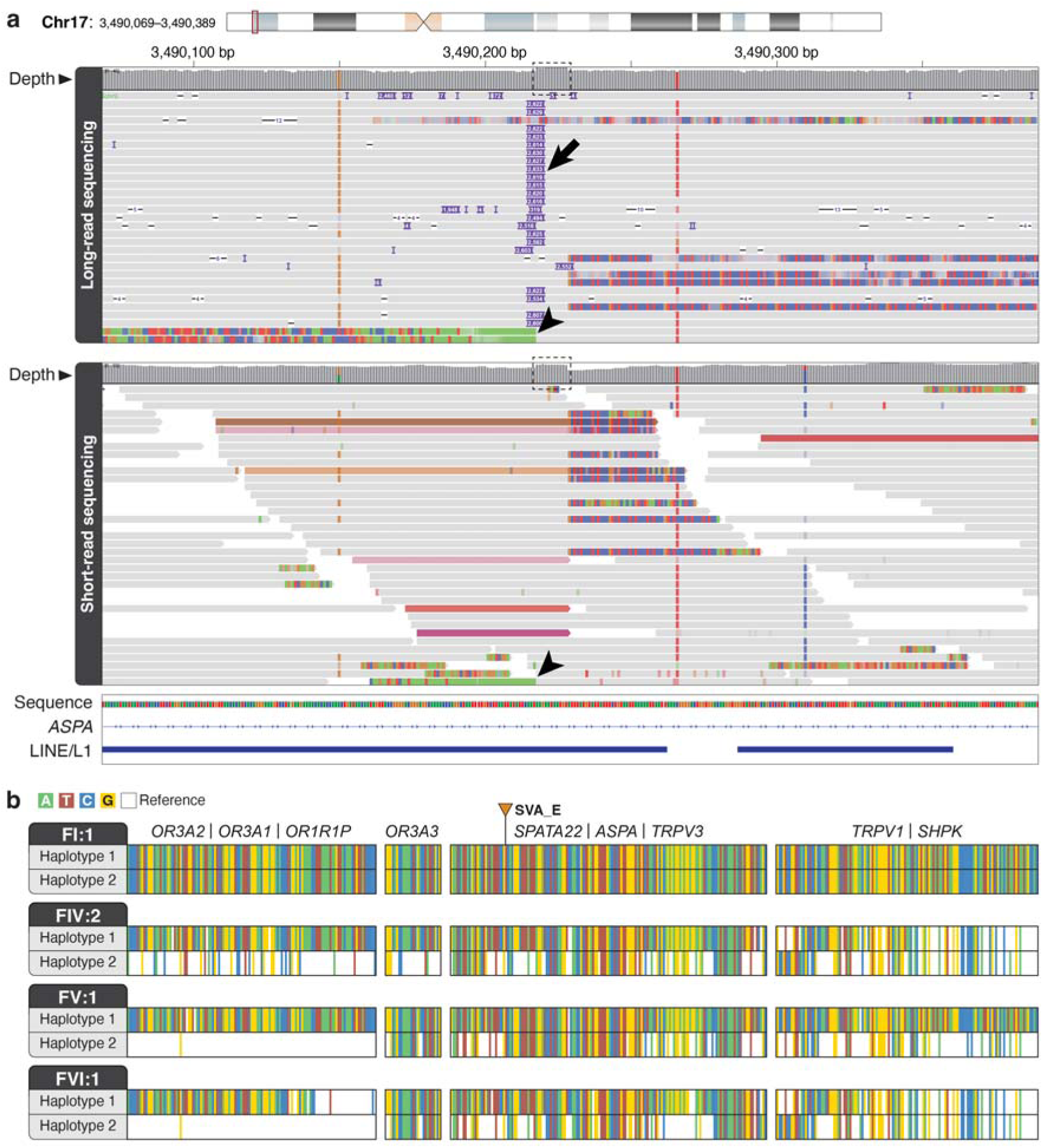
An SVA_E insertion in *ASPA* is associated with Canavan disease. **a.** IGV ^23^ view of LRS data (top) and srGS data (bottom) from individual FI:1 showing the SVA_E insertion (arrow). A target-site duplication is apparent in both the LRS and srGS data (dashed box). Colored reads in the srGS track indicate discordant read pairs, which occur when the mate of the read maps to another location in a genome. An increased number of these reads, as well as the soft-clipped sequences **(Fig. S1)** are seen at the site of a retrotransposon insertion. **b.** Analysis of phased heterozygous and homozygous SNVs for FI:1, FIV:2, FV:1, and FVI:1 demonstrates that SNVs from one haplotype in each affected individual within the region that includes *ASPA* are highly similar to SNVs from FI:1 suggesting a shared founder event. Only SNVs homozygous in FI:1 are plotted for FIV:2, FV:1 and FVI:1.

SVA insertions may disrupt transcription through various mechanisms^4^, including direct disruption of coding sequence or by causing aberrant splicing patterns that include exon skipping, activation of cryptic splice sites around the SVA insertion, and the inclusion of the SVA sequence in gene transcripts. To investigate the impact of this SVA_E insertion on gene function, we performed RNA-seq, with and without cycloheximide (CHX), on fibroblasts derived from a parental SVA carrier (family VI) and a proband (family VII). The untreated samples in both individuals lacked heterozygous variant calls that were present in the srGS data, indicating expression of only one allele (**Fig. 3b**). Both CHX-treated samples were observed to have soft clipping in sequencing reads spanning the exon 4 to intron 4 boundary that did not map to any *ASPA* genomic reference sequence but instead matched the sequence of the SVA_E present in intron 4 (**Fig. 3b**). This data indicated that the SVA_E insertion creates a novel splice acceptor site within intron 4 of *ASPA*, resulting in the disruption of canonical splicing and the addition of an SVA_E-containing exon (**Fig. 3a**). No heterozygous variants were observed downstream of the SVA_E-containing exon in the CHX treated samples, suggesting that transcription ends within the SVA_E insertion. The transcript incorporating the SVA_E was only observed in CHX-treated samples, indicating that the products from this allele are targeted for degradation. The RNA-seq data provided evidence that the deep intronic SVA insertion causes aberrant splicing that leads to transcript degradation and, consequently, loss of function of *ASPA*. Given that intronic retrotransposon insertions do not always cause aberrant splicing, performing RNA-seq or RT-PCR analyses is essential to confirm the functional consequences of such insertions.

**Fig. 3.**
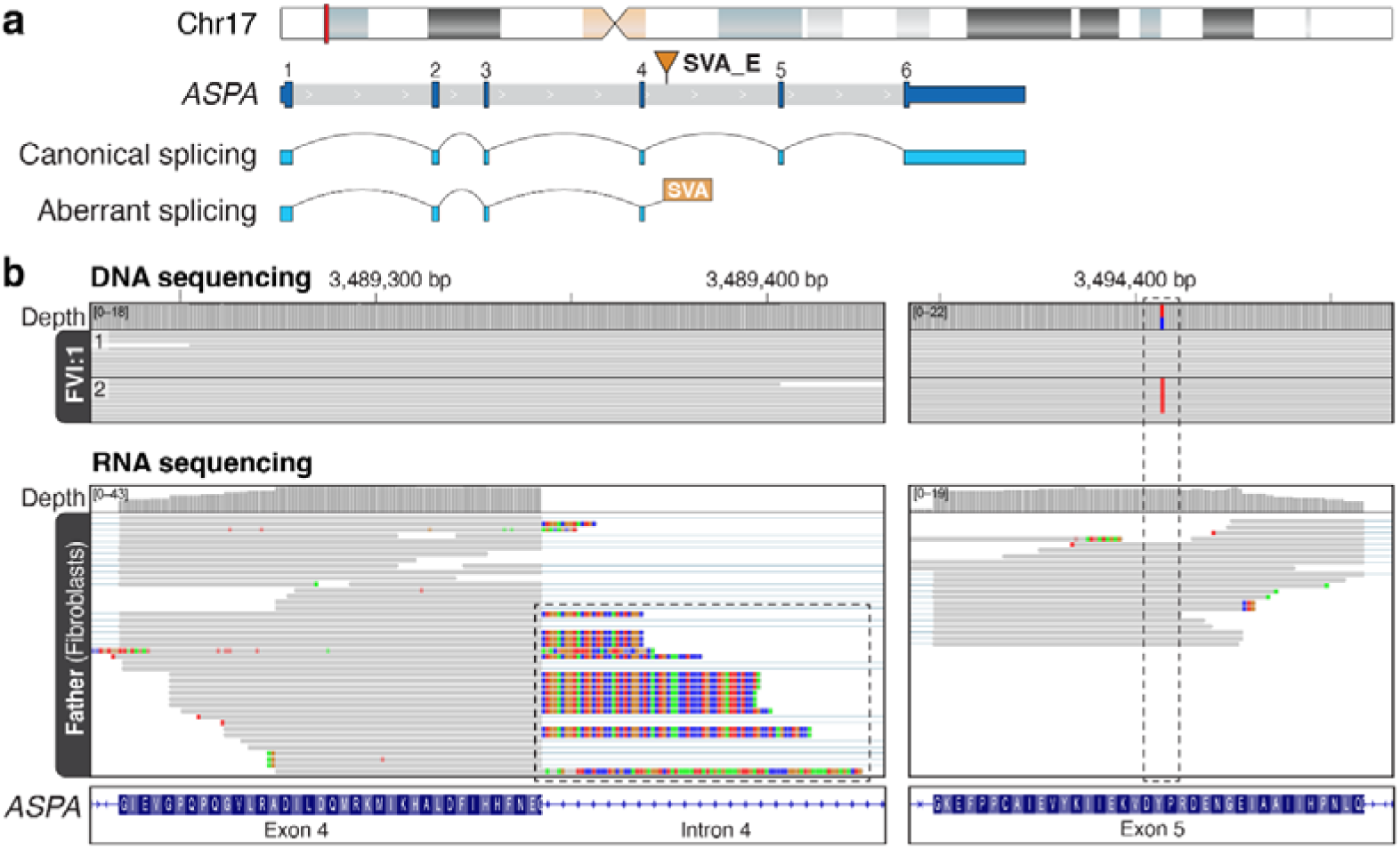
An intronic SVA_E insertion results in aberrant *ASPA* splicing. **a.** The position of *ASPA* on chr17 is shown, along with the gene structure and location of the SVA_E insertion in intron 4. Gray indicates intronic sequence, while blue represents exonic sequence, including the 5’ and 3’ UTRs. The structure of the canonical *ASPA* transcript is shown, along with an aberrant transcript that includes sequence from the SVA_E insertion. **b.** IGV screenshots showing phased DNA sequencing data from individual FVI:1, and RNA-seq of fibroblast-derived RNA from the father of FVI:1, who is heterozygous for the SVA_E insertion. Haplotype 2 from individual FVI:1 carries the SVA_E insertion and was paternally inherited. The fibroblast sample was treated with cycloheximide to inhibit nonsense-mediated decay. Soft-clipped sequence at the end of the RNA-seq data in exon 4 are observed (dashed box) which matches sequence from the SVA_E insertion, indicating splicing from exon 4 into the SVA_E. A heterozygous SNV in exon 5 is observed on haplotype 2—the same haplotype the SVA_E insertion is on—but is not observed in RNA-seq data (dashed box), suggesting that transcription ends within the SVA_E insertion.

The unexpected identification of the same SVA insertion in seven unrelated families suggested that either the insertion is a recurring event or that the variant is derived from a single past event. Analysis of heterozygous and homozygous SNVs within the target region (chr17:2,450,000-4,550,000) for individual FI:1 revealed no heterozygous SNVs, suggesting this is a large region of homozygosity in this individual **(Fig. 2b)**. Thus, we hypothesized that all individuals with the SVA_E insertion would have a similar pattern of SNVs on the haplotype with the insertion as FI:1. To evaluate this, we compared the homozygous SNVs from FI:1 with phased SNVs from FIV:1, FV:1, and FVI:1, who were all heterozygous for the SVA_E insertion by LRS. The analysis demonstrated that all three individuals carried SNVs with more than 99% identity to FI:1 in genes within the target region, confirming that this haplotype is indeed shared among these four individuals and that this disease-causing variant likely originated from a single insertion event **(Fig. 2b, Fig. S3)**. The presence of a shared haplotype could be used to infer the presence of the SVA_E insertion in individuals with a CD diagnosis in whom traditional testing did not identify the insertion.

We then evaluated the allele frequency of the SV insertion using gnomAD v4^5^. Out of approximately 126,000 haplotypes, 66 were heterozygous and zero homozygous for an SVA insertion at this position, yielding an allele frequency (AF) of 0.0005 (1/1,909 haplotypes carry the insertion) **(Table S2)**. In gnomAD, the SVA insertion was identified from srGS data using MELT^6^, which estimated the insertion length to be 461 bp. Due to the discrepancy in length between the SVA reported in gnomAD and our sequencing data, confirmation was obtained from gnomAD that the ends of the SVA_E insertion in our cohort matched the gnomAD data, providing evidence that they are the same variant. This variant is observed in all genetic ancestry groups in gnomAD except individuals of Middle Eastern descent, where fewer than 100 haplotypes are represented. Surprisingly, the most common previously reported pathogenic variant in *ASPA* (17-3494408-C-A) has an AF of 0.0003 (present in 1/3,314 haplotypes). The AF of the SVA_E insertion is below the maximum credible AF of 0.0013 for *ASPA* **(Fig. S4)**, meaning its AF is not high enough to raise concerns regarding the pathogenicity of the allele. This evidence suggests that the identified SVA_E is the most common disease-causing variant in *ASPA,* and it is not restricted to a specific genetic ancestry group.

While LRS was instrumental in defining the SVA_E insertion, analysis of short-read genome sequencing (srGS) data can detect the insertion through the presence of the tandem duplication site and the identification of split or discordant reads at the insertion breakpoints through IGV or by software such as MELT^6^ and xTea^7^ (**Fig. 2a**). The ability of TE software to detect the SVA_E insertion with srGS data was confirmed using xTea on sample FVI:1. The discordant read pairs exhibit a distinct pattern, with their mates aligning to multiple chromosomes due to the reads mapping to various SVA-containing regions in the human reference genome, as well as sequences from the SVA elements themselves. However, the repetitive nature and size of the SVA_E element necessitate LRS or PCR to fully characterize the insertion. This underscores a significant limitation of srGS: its inability to detect or fully resolve complex SVs, particularly those involving TEs.

Several reported individuals with clinical evidence of CD lack a definitive genetic diagnosis, with only one or no disease-causing variants identified in *ASPA*^8,9,10^. The number of molecularly unsolved CD cases is likely to be underreported since commercial genetic testing laboratories may not report single heterozygous pathogenic variants in autosomal recessive genes such as *ASPA*, and genetic confirmation of disease can be an exclusion criterion for case publication. Given this, revisiting previously unsolved CD cases to evaluate for this SVA_E insertion is vital. It is also important that genetic testing laboratories ensure they can detect this SVA_E insertion both in diagnostic testing and in prenatal carrier screening. This may require the use of LRS, PCR, or the application of TE-detecting software to identify the presence of mobile element insertions, such as retrotransposons, from short-read data. Alternatively, the described haplotype linked to the SVA_E insertion could be used to infer its presence. Finally, this finding highlights the importance of diagnostic orthogonal methods such as biochemical testing in cases where there is clinical suspicion for a specific condition such as CD, even in the context of nondiagnostic first-tier genetic testing.

This study highlights the identification of what may be the most common disease-causing variant in CD, which has been overlooked in 25 years of CD research. This finding is of great importance to the CD community, as it directly impacts families receiving a genetic diagnosis, as well as potential future therapeutic applications. In addition, this finding has significant implications for the broader rare disease community, as it highlights a significant blind spot in standard diagnostic pipelines that either miss or overlook these types of insertions. Specific software to detect TE insertions should be used in rare disease short-read data sets, or ideally, LRS deployed, especially where there is a strong clinical suspicion of a particular genetic disorder but traditional testing is nondiagnostic. As LRS technology continues to improve and becomes more accessible, it will likely help resolve the genetic etiology of other unsolved individuals and conditions. The growing number of studies analogous to the one presented here positions TE-related variants as an exciting avenue to explore in unsolved cases and shows that RNA-seq and LRS are powerful tools to solve them.

## Online Methods

### Patient ascertainment and clinical studies

Eight individuals from seven unrelated families of self-reported European-Uruguayan (FI:1, FII:1, FIII:1), European-American (FIV:1, FIV:2, FV:1), and European (FVI:1, FVII:1) ancestry were recruited and studied **(Table 1)**. Unless otherwise noted, patients were consented and enrolled under the Myelin Disorders Biorepository Project (MDBP) at The Children’s Hospital of Philadelphia (CHOP, IRB #14-011236), within the regulatory framework for the Global Leukodystrophy Initiative Clinical Trial Network (GLIA-CTN). The study was approved by the institutional ethics committees of the Children’s Hospital of Philadelphia, and written informed consent was obtained from all five families in accordance with the Declaration of Helsinki. For each individual enrolled, the medical records were collected from institutions where they received care. Family FVI was consented and enrolled under the Rare Diseases Now (RDNow): Genomic Diagnoses and Personalised Care for Children with Undiagnosed Rare Diseases program (HREC Reference Number HREC/67401/RCHM-2020) approved by the Royal Children’s Hospital Human Research Ethics Committee. Family FVII was consented under the Leukodystrophy Research Program at the Murdoch Children’s Research Institute (HREC#641943), approved by the Royal Children’s Hospital Human Research Ethics Committee. All individuals had a clinical diagnosis of CD based on biochemical testing and neuroimaging, but genetic testing had identified one or no known pathogenic or likely-pathogenic variants in the *ASPA* gene. Detailed clinical features, brain MRI imaging, family history, and clinical notes were reviewed by a group of pediatric neurologists and a genetic counselor. Brain MRIs were reviewed by experienced pediatric neuroradiologists.

**Table 1.**
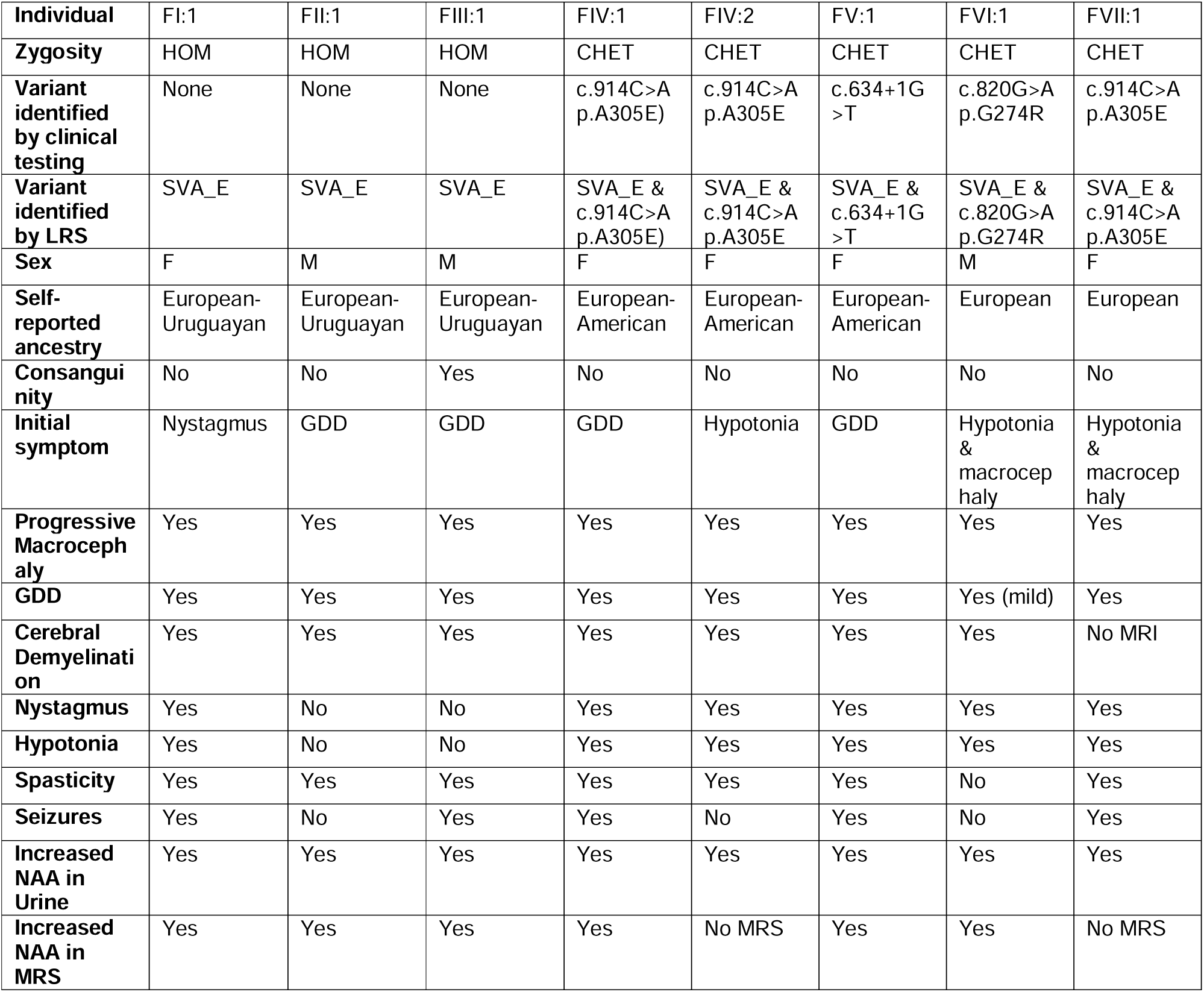
Summary of genotypes and phenotypes of individuals included in this report. HOM: homozygous, CHET: compound heterozygous, LRS: long-read sequencing, GDD: global developmental delay.

### Short-read sequencing of individual FI:1

SRS (2 x 150bp) was performed as a trio for patient FI:1 at the Children’s Hospital of Philadelphia High Throughput Sequencing core using a PCR-free protocol at a targeted mean coverage of >60x. The raw data (fastq) was aligned to the hg38 human reference genome and variant called (Single Nucleotide Variants or SNV, small insertion deletions or indels, Copy Number Variants or CNVs, and Regions of Homozygosity or ROH) using Illumina DRAGEN, v3.9.5^11^. The quality control protocol included the determination of sex from the genetic data and the estimation of kinship coefficients to confirm the relationships within the family. Sequence variants (SNVs and Indels) were annotated using the Variant Effect Predictor (VEP v106)^12^ and filtered against the gnomAD population database (v4)^5^ to remove variants that are observed in the general population (minor AF threshold at 0.5%). Sequence variants were further prioritized using an inhouse workflow to rank variants based on several factors, including prior evidence for pathogenicity using annotations from ClinVar^13^ and HGMD (Qiagen Inc., Germantown, MD)^14^ and several computational prediction scores, including CADD^15^, REVEL^16^, and SpliceAI^17^. CNVs were annotated using the AnnotSV^18^ and filtered for variants involving the protein-coding regions of the genome.

### Targeted long-read sequencing of affected individuals and their parents

Targeted LRS was performed on the Oxford Nanopore Technologies (ONT) platform for three unrelated individuals from our cohort. Briefly, HMW DNA was isolated from whole blood using the Puregene DNA Purification from a Blood kit (Qiagen) for patients FI:1 and FV:1. Residual DNA for FIV:2 was obtained from an outside laboratory. Extracted DNA was quantified on a Qubit Fluorometer (Invitrogen), and quality was assessed using a NanoDrop Spectrophotometer (ThermoFisher) and an Agilent Femto Pulse. The Short Read Eliminator kit (PacBio) was used to remove smaller fragments according to the manufacturer instructions for FV:1 and FIV:2. Libraries were prepared using the ONT Ligation Sequencing Kit (SQK-LSK114), loaded onto a R10.4.1 flow cell, and run using adaptive sampling on a PromethION 24. Targets were enriched using regions of approximately 2 Mbp surrounding the following genes: *HEPACAM, GALC, HEXA, GPRC5B, GCSH, ASPA, GFAP, AQP4, GCDH, MLC1, ARSA, AMT, HEXB, and GLDC*, and for approximately 200 kbp surrounding *FMR1* and *COL1A1* **(Table S1)**. Following sequencing, libraries were base called using Dorado version 0.5 (ONT) using the super accurate model with 5mCG and 5hmCG modifications. Run performance was evaluated using cramino (v0.14.1). Base called reads were aligned to GRCh38 using minimap2^19^ small variant calling and phasing was performed using Clair3^20^. SVs were called using Sniffles2^21^ and cuteSV^21, 22^. Reads were visualized using IGV^23^.

Targeted LRS was also performed on barcoded and pooled samples from the remaining affected individuals. Briefly, extracted DNA was quantified as above. Libraries were prepared using the ONT Native Barcoding kit (SQK-NBD114.24), the pool was loaded onto a R10.4.1 flow cell and run using adaptive sampling on a PromethION 24. The flow cell was washed and reloaded twice during a 66-hour run. Targets were enriched using the same regions as above **(Table S1)**. Following sequencing, libraries were base called using Dorado version 0.8.2 (ONT) using the super accurate model with 5mCG and 5hmCG modifications then demultiplexed using Dorado. Run performance was evaluated using cramino (v0.14.1)^24^. Base called reads were aligned to GRCh38 using minimap2^19^. Reads were visualized using IGV^23^.

### Illumina whole genome sequencing of families VI and VII

WGS data generation and clinical analysis were performed using diagnostically accredited methods by the Victorian Clinical Genetics Services (VCGS) in Melbourne, Australia. DNA was extracted manually from blood collected in EDTA vacutainers using the QIAamp DNA blood mini kit (QIAGEN). DNA quantity and quality were assessed using the Qubit dsDNA BR (broad-range) Assay kit (Thermo Fisher) and TapeStation genomic DNA kit (Agilent), respectively. Whole-genome DNA libraries were created using Nextera DNA Flex Library Prep kit/Illumina DNA prep kit (Illumina) followed by 2 × 150-bp paired-end DNA sequencing on a NovaSeq 6000 instrument (Illumina), variably using S2 or S4 flow cells. The targeted mean sequencing depth was 30×, with a minimum of 90% of bases sequenced to at least 10× for nuclear DNA (nDNA) and a minimum of 800× mean coverage for mitochondrial DNA (mtDNA).

### PacBio HiFi sequencing and alignment of individual VI:1

DNA for sequencing was homogenized using a Diagenode Megaruptor with the 3 DNAFluid + Kit (E07020001, Diagenode) using the following parameters: volume, 150 µL; speed, 40 and concentration 50 ng/µL. After homogenization, 3 ug of material was diluted in Low TE to a volume of 130 µL. Samples were sheared with the Megaruptor shearing kit (E07010003, Diagenode) using a speed of 30 or 31 depending on extraction fragment length, with the goal of recovering average fragment lengths of 15–24 kb. Clean-up and concentration of sheared material were performed with SMRT bell clean-up beads (102-158-300, PacBio) and eluted in a volume of 47 µL. Sheared average fragment lengths were determined by Femto pulse using the Genomic DNA 165kb Analysis Kit (FP-1002-0275, Agilent).

SMRTbell libraries were prepared using SMRTbell prep kit 3.0 (102-141-700, PacBio) following the standard procedure and performing the unique barcoding of each sample with the SMRTbell adapter index plate 96A (102-009-200, PacBio). Size selection was performed using the AMPure PB beads size selection kit (102-182-500, PacBio) at a ratio of 2.9x (i.e., 50 µL of sample: 145 µL 35% beads). Size determination of the final SMRT library was performed using the Femto pulse as described above. SMRT Library average fragment lengths ranged between 12.895 and 26.001 Kb, and final SMRT libraries were diluted to below 60 ng/µL before loading. Samples were sequenced with a movie time of 30 hours and using SMRT Link version 13.1.0.221970, chemistry bundle: 13.1.0.217683, and parameter version 13.1.0. The following PacBio products were used for the sequencing reaction: Revio sequencing plate (02-587-400) and Revio Polymerase kit (102-739-100) and Revio SMRT cell tray (102-202-200). The On Plate Loading Concentration was set to 250 pM for all sequencing runs. Any sample that did not generate 90 Gb using 1 SMRT cell was repeated on pooled runs by combining SMRT libraries in “top-up” runs.

After sequencing, HiFi FASTQ files were aligned to the hg38 reference genome using minimap2 (v2.14-r883)^19^. SNVs and indels were called using Clair3 (v1.0.4)^20^ and phased using WhatsHap (v2.1)^25^. SVs (including CNVs) were called with Sniffles2^21^ (v2.07). SNVs and indels were functionally annotated using Variant Effect Predictor (VEP)^26^ (v110), and SVs were annotated using AnnotSV (v3.3.4)^18^.

### RNA Sequencing

Patient fibroblast cell lines were established, grown to 7×10^5^ Cell/mL, then treated with or without cycloheximide for 22–24 hours. Cells were centrifuged, and the resulting cell pellet was washed with PBS. Total RNA was extracted from the cell pellets, and the Illumina Stranded mRNA protocol was used for library prep, which was sequenced to a depth of 80 million reads. RNA sequencing data was mapped to the hg38 reference genome using STAR v2.7.3a^27^. The 2-pass method within STAR was utilized with gencode.v35 gene annotations to enhancer mapping and the detection of unique splicing events. Data quality was assessed with FastQC, and data was visualized using IGV^23^.

### Establishing the maximum credible allele frequency for *ASPA*

The maximum credible AF was calculated using an established statistically robust framework for assessing whether a variant is “too common” to be causative for a Mendelian disorder of interest^28^ **(Fig. S4a)**. To calculate the maximum allele contribution in *ASPA,* we used data from gnomAD (v4)^5^ **(Table S3)**. The allele count (467) of the most common previously described pathogenic allele (17-3494408-C-A) was divided by the total number of pathogenic or likely pathogenic alleles (1021). The prevalence of CD used for the calculation was 1 in 10,000, penetrance was determined to be one since CD is fully penetrant, and genetic heterogeneity was also one as it is a monogenic disease. The Frequency Filter App was used to perform the calculation **(Fig. S4b)**.

## Supplementary Material

**Table S1 | Target regions for adaptive sampling using GRCh38 coordinates.**

Excel File

**Table S2 | List and allele frequencies of pathogenic or likely pathogenic *ASPA* variants from gnomAD v4 sorted by allele frequency.**

Excel File

**Fig. S1.**
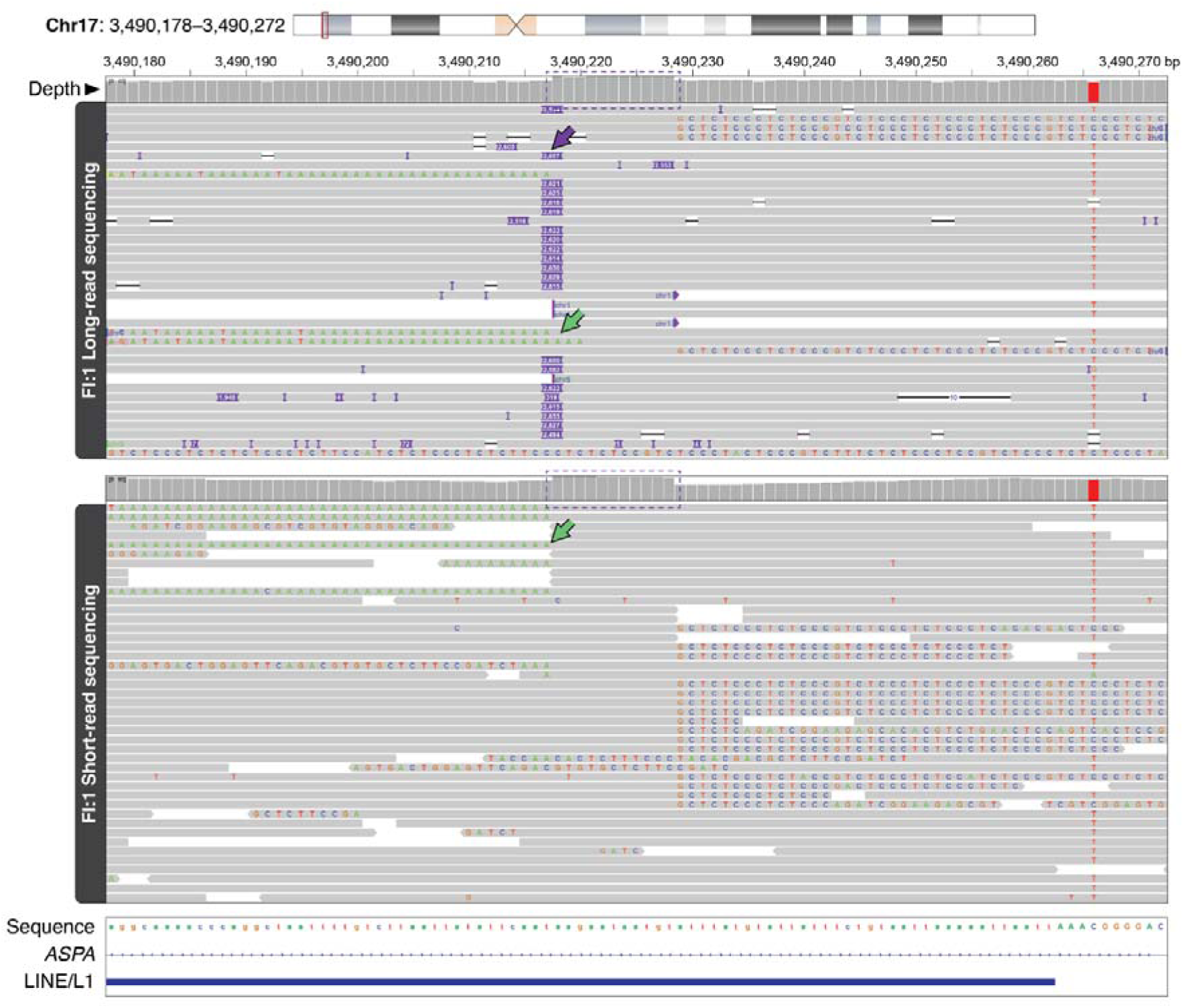
IGV view of SVA_E insertion with clipped reads shown. IGV view of LRS and srGS data from individual FI:1 with clipped reads shown. The clipped reads contain poly-A sequence located at the end of the SVA_E insertion on the left side of the target site duplication, and sequence from the start of the SVA_E insertion to the right.

**Fig. S2.**
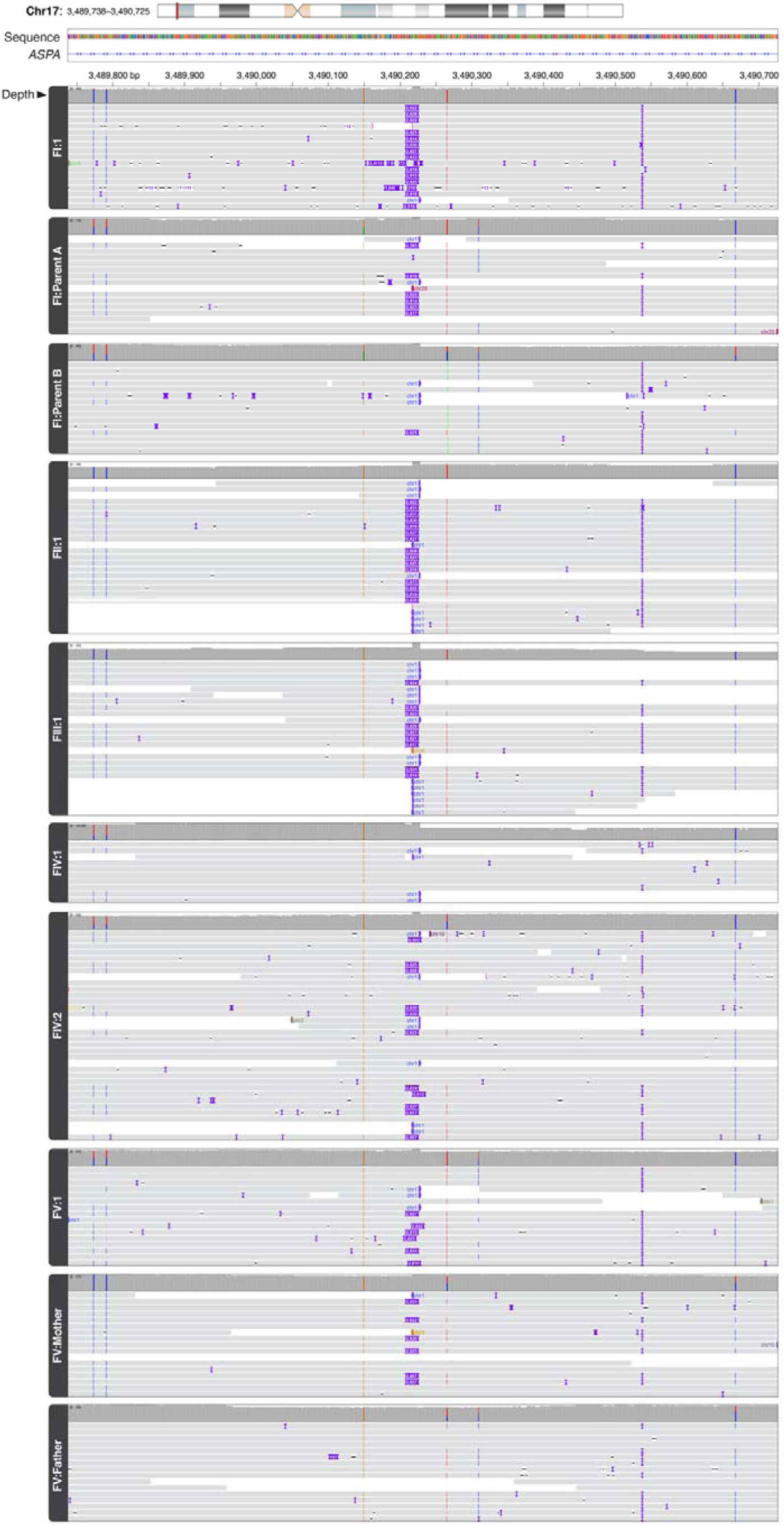
Long-read sequencing of families I–V. IGV views of a 989-bp region (chr17:3,489,738-3,490,725) for individuals from five of the seven families reported here. Whole-genome LRS was performed for the proband from family FI, while targeted LRS was performed for FIV:1 and FV:1. A single adaptive sampling run after barcoded library prep was performed for all other individuals shown here. Sequencing confirms that the parents from F1 are heterozygous carriers of the SVA_E insertion, that both affected siblings from FIV are heterozygous for the SVA_E insertion (no insertion is shown because of low coverage, but four reads are clipped at the insertion position), and that the mother from FV is heterozygous for the SVA_E insertion.

**Fig. S3.**
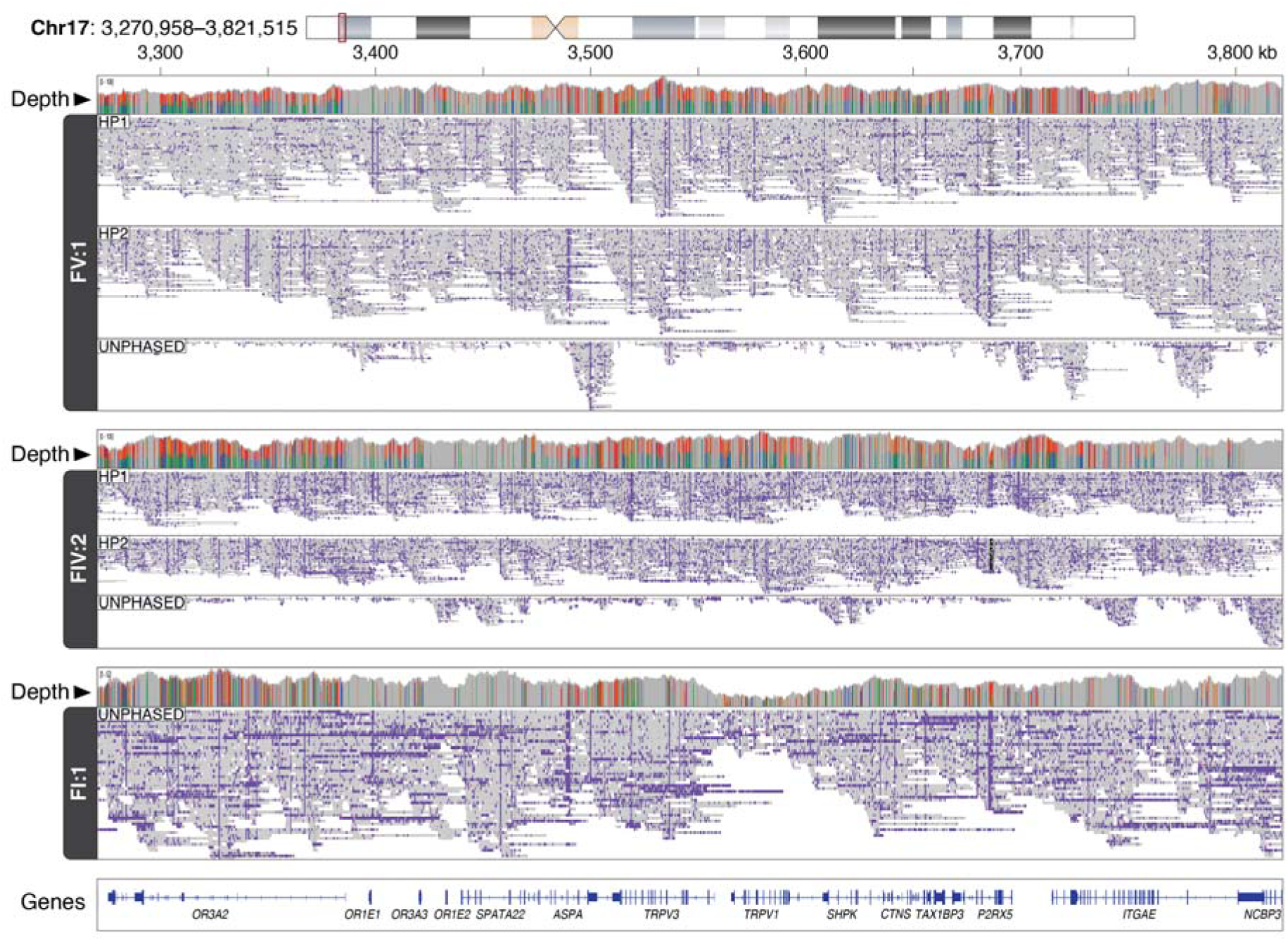
Identification SVA_E insertion in intron 4 of *ASPA*. IGV view of a 550-kbp region (chr17:3,270,958-3,821,515, GRCh38) surrounding *ASPA* demonstrates phasing in heterozygous individuals FV:1 and FIV:2. Individual FI:1 is not phased as all SNVs are homozygous in this region.

**Fig. S4.**
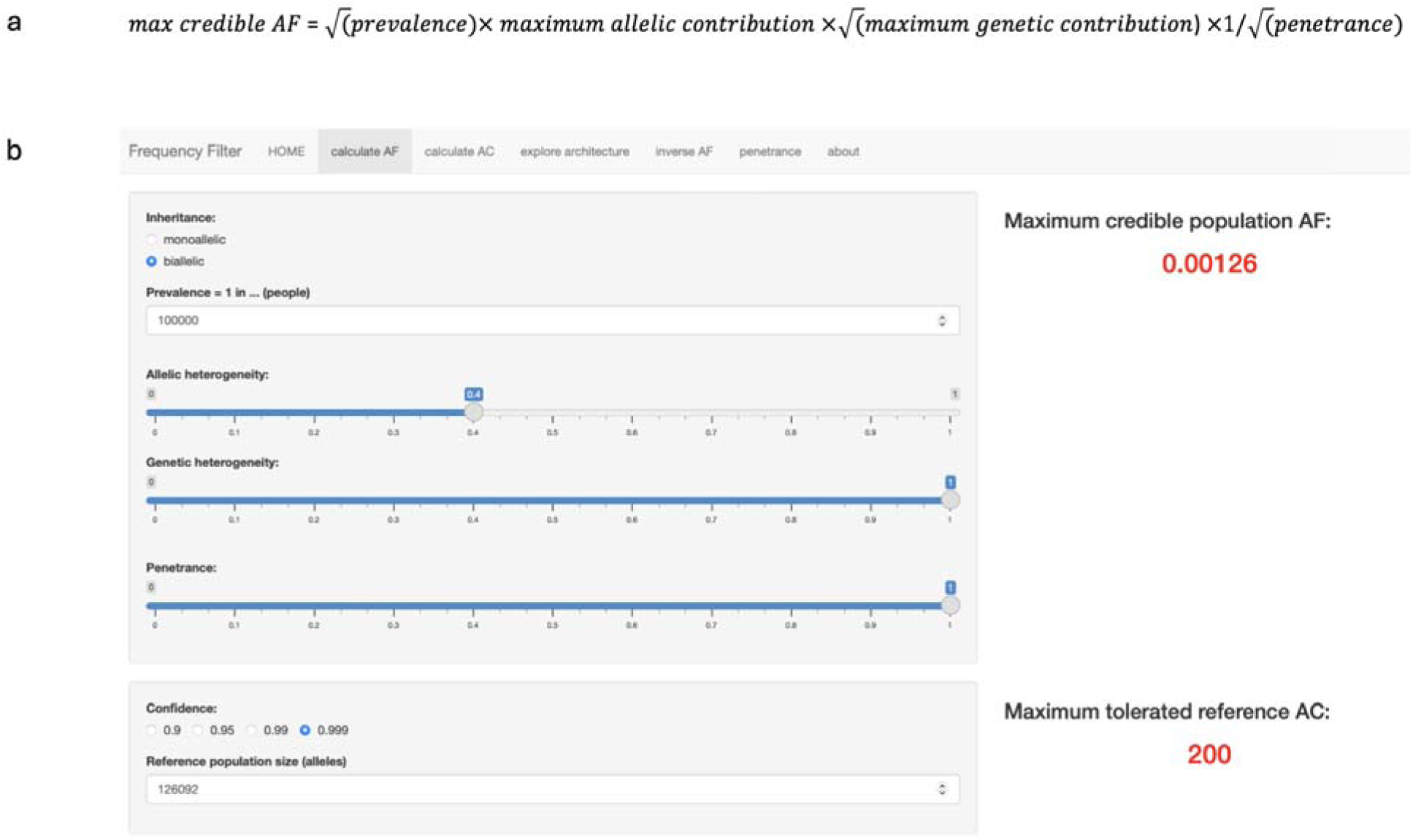
Maximum credible allele frequency for *ASPA*. **a.** The formula used to calculate the upper bound for the maximum credible AF of an individual causative variant in the *ASPA* gene^28^. **b.** Screenshot of The Frequency Filter App showing the parameters used to calculate the upper bound for the maximum credible AF of an individual causative variant in the *ASPA* gene, and the resulting values.

## Case Reports

We identified eight probands from seven unrelated families with a clinical diagnosis of Canavan disease based on biochemical testing and clinical presentation who did not have a precise or complete molecular diagnosis **(Fig. 1a)**. For FI:1, FII:1, and FIII:1, previous commercial and/or research-based genetic testing did not report any variants in *ASPA*. For FIV:1, FIV:2, FV:1, FVI:1, and FVII:1 previous genetic testing identified a single pathogenic variant in *ASPA*, but no second variant.

### Individual FI:1

A female proband born to a non-consanguineous couple with no significant family history. Initial MRI of the brain showed diffuse abnormality of the white matter affecting the subcortical and deep white matter as well as brainstem and cerebellum **(Fig. 1a)**. Spectroscopy showed an elevated NAA peak. A Leukodystrophy gene panel (lnvitae) did not report any variants in *ASPA*. Subsequent clinical SRS was non-diagnostic. Markedly elevated excretion of NAA was identified by urine organic acid analysis (1582.0 mmol/mol creatinine). Clinical findings are remarkable for progressive macrocephaly, global developmental delay, seizures, spasticity, constipation/reflux, and G-tube dependency. Most recent neuroimaging findings are remarkable for leukodystrophy with involvement of the entire white matter, globus pallidus, putamen, brainstem, and cerebellum, consistent with Canavan disease **(Fig. 1b)**.

### Individual FII:1

A male presented with global developmental delay **(Fig. 1a)**. Clinical findings were remarkable for global developmental delay with incomplete head support, seizures, dystonic-spastic quadriparesis, and progressive macrocephaly. Initial brain MRI showed increased signal in T2 and FLAIR sequences and low signal in T1 at the level of the diffuse white matter, supra- and infratentorial, thalami and pallidum respecting the internal capsules, caudate, putamen and corpus callosum **(Fig. 1b)**. Spectroscopy showed an elevated NAA peak **(Fig. 1c)**. Clinical Sanger sequencing and MLPA analysis of *ASPA* were performed on a clinical basis and did not identify any pathogenic variants. Markedly elevated excretion of NAA was observed by urine organic acid analysis (analyzed qualitatively by gas chromatography-mass spectrometry).

### Individual FIII:1

A male proband born to a non-consanguineous couple with no significant family history **(Fig. 1a)**, presented with global developmental delay. Clinical findings were remarkable for progressive macrocephaly, severe global developmental delay with incomplete head support, irritability, and increased muscle tone, and death due to disease progression. MRI of the brain revealed increased signal in T2 and FLAIR in the supratentorial subcortical, deep and periventricular white matter, thalami and pallidum, cerebral peduncles, posterior sector of the pons, medulla oblongata, dentate nuclei, and medial cerebellar peduncles **(Fig. 1b)**. Spectroscopy revealed an elevated NAA peak **(Fig. 1c)**. Clinical Sanger sequencing and MLPA analysis of *ASPA* were performed on a clinical basis and did not identify any pathogenic variants. SRS was performed on a research basis and did not identify any variants of interest. Markedly elevated excretion of NAA was identified in urine organic acid analysis (analyzed qualitatively by gas chromatography-mass spectrometry).

### Individual FIV:1

A female presented after her older sister (FIV:2) was diagnosed with Canavan disease **(Fig. 1a)**. Clinical findings were remarkable for progressive macrocephaly, global developmental delay, nystagmus, irritability, seizures, and increased muscle tone. MRI of the brain showed restricted diffusion in the centrum semiovale symmetrically along the cortical spinal tracts and in the dorsal brainstem as well as cerebellar white matter symmetrically **(Fig. 1b)**. Spectroscopy showed an elevated NAA peak **(Fig. 1c)**. Rapid whole genome sequencing identified a single heterozygous pathogenic variant in *ASPA* c.914C>A (p.Ala305Glu) that was not maternally inherited (father did not submit a sample). Urine organic acid analysis revealed markedly elevated excretion of NAA.

### Individual FIV:2

A female presented with a history of decreased fetal movement and decreased muscle tone at birth **(Fig. 1a)**. Clinical findings were remarkable for developmental delay, congenital nystagmus, spasticity, metabolic acidosis, seizures, constipation, progressive macrocephaly, hip subluxation, and neuromuscular scoliosis. Brain MRI at was noted to have elevated T2 signal on FLAIR in deep white matter with commensurate low signal on T1 and diffusion restriction of the same areas. Subsequent MRIs showed diffuse leptomeningeal enhancement, diffuse bilateral white matter T2 hyperintensity affecting the subcortical U fibers, and diffuse atrophy of the brain stem **(Fig. 1b)**. No Magnetic Resonance Spectroscopy was performed. Markedly elevated excretion of NAA identified in urine organic acid analysis (2712.76 mmol/mol creatinine). Targeted *ASPA* gene sequencing identified a heterozygous c.914C>A (p.Ala305Glu) variant classified as pathogenic.

### Individual FV:1

A female presented due to deficits with visual tracking and head control **(Fig. 1a)**,. Clinical findings were remarkable for macrocephaly, encephalopathy, hypotonia, nystagmus, exaggerated startle, seizures, developmental delay, chronic constipation, and hepatosplenomegaly. MRI of the brain remarkable for T2 signal abnormalities throughout the supratentorial white matter, with additional T2 signal abnormalities affecting the globus pallidus, thalamus, internal capsule, dorsal brainstem, cerebellar white matter with enlarged perivascular spaces **(Fig. 1b)**. Spectroscopy showed an elevated NAA peak **(Fig. 1c).** Markedly elevated excretion of NAA identified in urine organic acid analysis (414mg/g creatinine). Exome sequencing was initially negative, however after reanalysis with updated biochemical results, the updated report included a heterozygous paternally inherited pathogenic splice site variant *ASPA* c.634+1G>T.

### Individual FVI:1

A male proband presented with head lag, progressive macrocephaly (98^th^ percentile) and hypotonia **(Fig. 1a)**. An ultrasound at 32 weeks gestation showed mild ventriculomegaly (9.6mm), that resolved on follow-up. MRI brain demonstrated significant decrease in T1 and marked increase in T2 signal within supratentorial white matter, involving subcortical U fibers and sparing periventricular regions. Increased T2 signal was also seen within the thalami, globi palladi, midbrain, dorsal pons, medulla, cerebellar white matter, and dentate nuclei, with sparing of caudates and putamina. Spectroscopy revealed an elevated NAA peak within the basal ganglia (NAA:Cr of 2.29). Urinalysis identified a marked increase in N-acetylaspartic acid. Clinical findings were remarkable for horizontal nystagmus, hypotonia, progressive macrocephaly, and mild global developmental delay. There was no spasticity, irritability, sleep disturbance or regression. He was cruising on furniture and walking confidently in a walking frame. He had two-word phrases and good receptive language. He was developing a pincer grip and feeding himself. Targeted NGS of *ASPA* identified a single pathogenic variant: c.820G>A; (p.Gly271Arg), subsequently confirmed to be maternally inherited. He also has a maternally inherited chromosome 6q14.1 duplication of uncertain significance identified on chromosome microarray.

### Individual FVII:1

A female presented with hypotonia, developmental delay and macrocephaly **(Fig. 1a)**. Urine organic acid analysis revealed markedly elevated excretion of NAA consistent with a diagnosis of Canavan disease. *ASPA* sequencing identified a single heterozygous pathogenic variant in *ASPA* c.914C>A (p.Ala305Glu). There was no indication for brain MRI/MRS. Clinical findings include spastic quadriplegia with central hypotonia. She can grip light objects and use a large switch button to turn a toy on. She is non-verbal and can communicate yes/no and simple choices with blinking, vocalization and eye gaze. She has a severe cortical vision impairment with light sensitivity and intermittent nystagmus. She is tube feeding dependent and being treated for severe GERD, epilepsy, constipation, and saliva management. She had a bilateral pelvic/femoral osteotomy for hip dysplasia.

## Acknowledgements

We thank Angela Miller for help with manuscript and figure preparation.

## Conflict Of Interest

MPGZ and JG have received travel support from ONT. AV receives grant and in-kind support for translational research without personal compensation from Affinia, Biogen, Boehringer Ingelheim, Eli Lilly, Illumina, Ionis, Homology, Myrtelle, Orchard therapeutics, Passage Bio, Sana, Sanofi, Synaptixbio, and Takeda. DEM holds stock options in MyOme and Basis Genetics, is on a scientific advisory board at ONT and Basis Genetics, is engaged in a research agreement with ONT, and has received travel support from ONT and PacBio.

## Funding

The Rare Disease Flagship acknowledges financial support from the Royal Children’s Hospital Foundation and the Murdoch Children’s Research Institute, Melbourne, Australia. The research conducted at the Murdoch Children’s Research Institute was supported by the Victorian Government’s Operational Infrastructure Support Program. Massimo’s Mission acknowledges financial support from the Australian Government Department of Health and Aged Care (EPCD000034). AV is funded by the RDCRN (U54TR002823) and by 5U24NS131172. DEM is supported by the National Institutes of Health through the NIH Director’s Early Independence Award, DP5-OD033357.

## Data Availability

Additional data that support the findings of this study are available. De-identified genomic and associated data from this study are available on request from the corresponding author through formal data sharing agreements. All variants reported in this study have been deposited in ClinVar (SCV005407762).

The web resources used are:

ClinVar: https://www.ncbi.nlm.nih.gov/clinvar

gnomAD: https://gnomad.broadinstitute.org

IGV: https://software.broadinstitute.org/software/igv

NCBI RefSeq: https://www.ncbi.nlm.nih.gov/refseq

OMIM: https://omim.org

Frequency Filter: https://cardiodb.org/allelefrequencyapp/

UCSC: https://genome.ucsc.edu/

## Notes

### Funding Statement

The Rare Disease Flagship acknowledges financial support from the Royal Childrens Hospital Foundation and the Murdoch Childrens Research Institute, Melbourne, Australia. The research conducted at the Murdoch Childrens Research Institute was supported by the Victorian Government's Operational Infrastructure Support Program. Massimos Mission acknowledges financial support from the Australian Government Department of Health and Aged Care (EPCD000034). AV is funded by the RDCRN (U54TR002823) and by 5U24NS131172. DEM is supported by NIH grant DP5OD033357.

### Author Declarations

Ethics committee/IRB of The Children's Hospital of Philadelphia gave ethical approval for this work. Royal Children's Hospital Human Research Ethics Committee gave ethical approval for this work.

